# A genetically-informed study disentangling the relationships between tobacco smoking, cannabis use, alcohol consumption, substance use disorders and respiratory infections, including COVID-19

**DOI:** 10.1101/2021.02.11.21251581

**Authors:** Daniel B. Rosoff, Joyce Yoo, Falk W. Lohoff

**Affiliations:** Section on Clinical Genomics and Experimental Therapeutics, National Institute on Alcohol Abuse and Alcoholism, National Institutes of Health, Bethesda, MD, USA; NIH-Oxford-Cambridge Scholars Program; Nuffield Department of Population Health, University of Oxford, UK

## Abstract

**Background:** Observational studies suggest smoking, cannabis use, alcohol consumption, cannabis use, and substance use disorders (SUDs) may play a role in the susceptibility for respiratory infections and disease, including coronavirus 2019 (COVID-2019). However, causal inference is challenging due to comorbid substance use.

**Methods:** Using genome-wide association study data of European ancestry (data from >1.7 million individuals), we performed single-variable and multivariable Mendelian randomization to evaluate relationships between smoking, cannabis use, alcohol consumption, SUDs, and respiratory infections.

**Results:** Genetically predicted lifetime smoking was found to be associated with increased risk for hospitalized COVID-19 (odds ratio (OR)=4.039, 95% CI 2.335-6.985, *P*-value=5.93×10^−7^) and very severe hospitalized COVID-19 (OR=3.091, 95% CI, 1.883-5.092, *P*-value=8.40×10^−6^). Genetically predicted lifetime smoking was also associated with increased risk pneumoniae (OR=1.589, 95% CI, 1.214-2.078, *P*-value=7.33×10^−4^), lower respiratory infections (OR=2.303, 95% CI, 1.713-3.097, *P*-value=3.40×10^−8^), and several others. Genetically predicted cannabis use disorder (CUD) was associated with increased bronchitis risk (OR=1.078, 95% CI, 1.020-1.128, *P*-value=0.007).

**Conclusions:** We provide strong genetic evidence showing smoking increases the risk for respiratory infections and diseases even after accounting for other substance use and abuse. Additionally, we provide find CUD may increase the risk for bronchitis, which taken together, may guide future research SUDs and respiratory outcomes.

## INTRODUCTION

Since the first reported cases in Wuhan, China in December 2019,^1^ coronavirus disease 2019 (COVID-19) has subsequently affected more than 200 countries and continues to be a global pandemic of substantial worldwide morbidity and mortality.^2,3^ More broadly, upper and lower respiratory infections (URIs and LRIs, respectively), and other respiratory diseases (i.e., asthma, chronic obstructive pulmonary disease (COPD), etc.) are leading causes of yearly worldwide morbidity and mortality.^4,5^ For example, the Global Burden of Disease Study estimated that LRIs caused more than two million deaths globally in 2016,^4^ while approximately 2.3 million people died from COPD in 2015.^5^ Respiratory infection and diseases are also a large economic burden: URIs result in more than 40 million missed days of school and work per year.^6^

Substance use (tobacco smoking, cannabis use, and alcohol consumption) are risk factors linked with adverse lung and respiratory outcomes.^7-9^ For example, observational data has shown chronic heavy alcohol consumption to be associated with increased risk for pneumonia^7^ and acute respiratory distress syndrome,^10^ while cannabis smoke has been shown to contain many of the same toxins and irritants as smoke derived from tobacco,^11^ but may differ than tobacco in its association with bronchitis and other respiratory infections.^12^ In addition, it has been suggested that chronic alcohol abuse may compromise the ability of immune cells to destroy bacteria in the lungs, which may result in an increased vulnerability to respiratory infections like pneumonia and tuberculosis.^13^

Paralleling the COVID-19 pandemic have been increases in substance use,^14^ which combined with data showing approximately 10.8% of US adults have a substance use disorder (SUD),^15^ and recent work using electronic health records (EHRs) to show that individuals with a SUD are at increased risk for COVID-19^16^ suggest identifying potential causal relationships between substance use, SUD and respiratory infectious disease would have substantial public health benefit.

However, observational studies cannot be used to reliably identify causality due to limitations such as residual confounding, and reverse causality.^17^ For example, outcomes reached from observational studies may be subject to unmeasured confounders like comorbid disorders or underlying genetic differences that may lead to biased estimates, and consequently, may not reflect true causal relationships.^18,19^ While, randomized controlled trials (RCTs) are considered the “gold standard”, RCTs can be both unethical and impractical:^20,21^ Constructing an RCT to examine the effect of substance use on respiratory infection risk may be complicated by other existing comorbidities.

Mendelian randomization (MR) is a genetic approach that uses genetic variants as instrumental variables to explore causal relations between exposures (e.g. alcohol consumption, tobacco smoking, cannabis use) and health outcomes (e.g. respiratory infections and diseases). This technique takes publicly available genome wide association studies to screen for suitable genetic instrumental variables, which allows researchers to perform MR studies without the need to recruit new patients.^22^ Because germline variants are randomly assorted at meiosis, MR may be considered conceptually equivalent to RCTs, though a more naturalized version.^19,22^ More specifically, given genetic instruments cannot be influenced by other confounders (i.e., lifestyle, or environmental factors), MR studies, are in theory, less susceptible to confounding or reverse causality than traditional observational studies.^23^ Therefore, MR are an important analytical approach to strengthen causal inference when RCTs are challenging due to methodological or ethical constraints.^24^

Given the potential for confounding and limited causal inference derived from observational data, we used large, publicly available genome-wide association study (GWAS) data and two-sample MR methods to evaluate the relationships between substance use, substance use disorders (CUD and alcohol use disorder (AUD)) and respiratory infection and disease outcomes.

## METHODS

### Data sources and genetic instruments

Summary-level data for both modifiable risk factor instrument and infectious disease outcome data were derived from publicly available GWASs in populations of predominantly European ancestry (**Figure 1; Table 1 in the Supplement**). All GWASs have existing ethical permissions from their respective institutional review boards and include participant informed consent and included rigorous quality control.

**Table 1.** Single Variable MR Results of the Genetic Liability of Alcohol, Cannabis and Lifetime Smoking Exposures on COVID-19 Outcomes

| Outcome | Exposure | N SNPs | Odds Ratio | CI Lower Bound | CI Upper Bound | P-value |
| --- | --- | --- | --- | --- | --- | --- |
| Hospitalized v not hospitalized | Lifetime smoking | 91 | 1.710 | 0.689 | 4.240 | 0.247 |
|  | Cannabis use | 26 | 1.104 | 0.888 | 1.371 | 0.373 |
|  | Cannabis use disorder | 38 | 0.991 | 0.744 | 1.320 | 0.949 |
|  | Drinks per week | 71 | 1.229 | 0.490 | 3.079 | 0.660 |
|  | Alcohol use disorder | 10 | 0.897 | 0.700 | 1.151 | 0.393 |
| Hospitalized v population (European) | Lifetime smoking | 91 | 4.039 | 2.335 | 6.985 | **5.93E-07** |
|  | Cannabis use | 25 | 1.079 | 0.929 | 1.252 | 0.320 |
|  | Cannabis use disorder | 37 | 1.032 | 0.829 | 1.285 | 0.780 |
|  | Drinks per week | 70 | 0.906 | 0.473 | 1.733 | 0.765 |
|  | Alcohol use disorder | 9 | 0.931 | 0.779 | 1.113 | 0.434 |
| Very severe hospitalized v not hospitalized | Lifetime smoking | 91 | 3.097 | 1.883 | 5.092 | **8.40E-06** |
|  | Cannabis use | 25 | 0.937 | 0.454 | 1.933 | 0.860 |
|  | Cannabis use disorder | 38 | 1.010 | 0.862 | 1.183 | 0.901 |
|  | Drinks per week | 71 | 0.882 | 0.505 | 1.540 | 0.658 |
|  | Alcohol use disorder | 10 | 0.921 | 0.789 | 1.076 | 0.302 |
| COVID-19 v self-reported negative | Lifetime smoking | 91 | 0.852 | 0.596 | 1.217 | 0.379 |
|  | Cannabis use | 25 | 1.031 | 0.957 | 1.112 | 0.420 |
|  | Cannabis use disorder | 38 | 0.940 | 0.822 | 1.076 | 0.370 |
|  | Drinks per week | 70 | 0.870 | 0.608 | 1.245 | 0.447 |
|  | Alcohol use disorder | 10 | 1.128 | 1.039 | 1.226 | **0.004** |
| COVID-19 v population (European) | Lifetime smoking | 91 | 1.187 | 0.856 | 1.647 | 0.304 |
|  | Cannabis use | 25 | 1.019 | 0.945 | 1.098 | 0.627 |
|  | Cannabis use disorder | 38 | 0.964 | 0.843 | 1.101 | 0.588 |
|  | Drinks per week | 70 | 0.799 | 0.554 | 1.154 | 0.232 |
|  | Alcohol use disorder | 10 | 1.102 | 0.991 | 1.225 | 0.073 |
| COVID-19 v population | Lifetime smoking | 91 | 1.144 | 0.845 | 1.550 | 0.383 |
|  | Cannabis use | 25 | 1.046 | 0.983 | 1.114 | 0.154 |
|  | Cannabis use disorder | 38 | 0.974 | 0.870 | 1.090 | 0.644 |
|  | Drinks per week | 71 | 0.818 | 0.579 | 1.156 | 0.254 |
|  | Alcohol use disorder | 10 | 1.061 | 0.972 | 1.158 | 0.188 |
| Predicted COVID-19 v self-reported negative | Lifetime smoking | 91 | 0.887 | 0.476 | 1.653 | 0.705 |
|  | Cannabis use | 24 | 1.093 | 0.963 | 1.241 | 0.171 |
|  | Cannabis use disorder | 37 | 1.036 | 0.802 | 1.339 | 0.787 |
|  | Drinks per week | 71 | 0.664 | 0.332 | 1.327 | 0.246 |
|  | Alcohol use disorder | 10 | 1.126 | 0.965 | 1.314 | 0.131 |

Abbreviations: MR, mendelian randomization; GWAS, genome wide association study; N SNPs, number of single-nucleotide polymorphism (genetic instruments); OR, odds ratio; CI, confidence interval.
a Results from two sample SVMR inverse-variance weighted MR analysis; outliers identified by MR PRESSO tool were removed; estimated associations reported as odds ratios with 95% confidence intervals. Boldface indicates conventional statistical significance ( $P < 0.05$ ).
b Genetic instruments selected from 5 GWASs, selection threshold $P < 5 \times 10^{-8}$ or $P < 5 \times 10^{-6}$ , clumped at linkage disequilibrium (LD) $r^2 = .001$ (10 000 kilobase pair window); N SNPs differs across outcomes depending on number of genetic instruments found in outcome GWASs.

**Fig 1.**
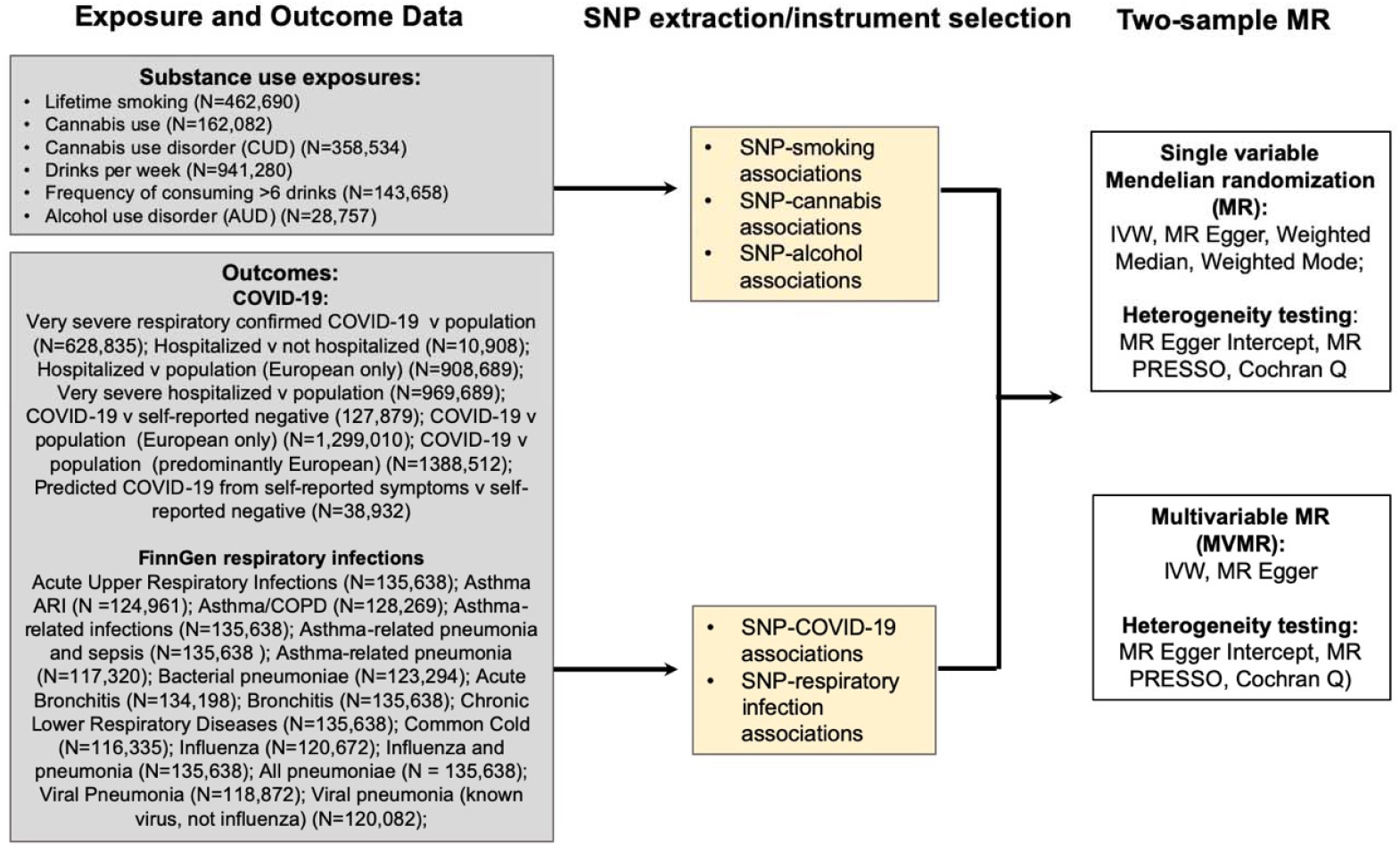
Study overview. Abbreviations: SNP: single nucleotide polymorphism; COVID-19: coronavirus disease 2019; IVW, Inverse Variance Weighted MR; SVMR; single variable Mendelian randomization; MVMR: multivariable Mendelian randomization

**Fig 2.**
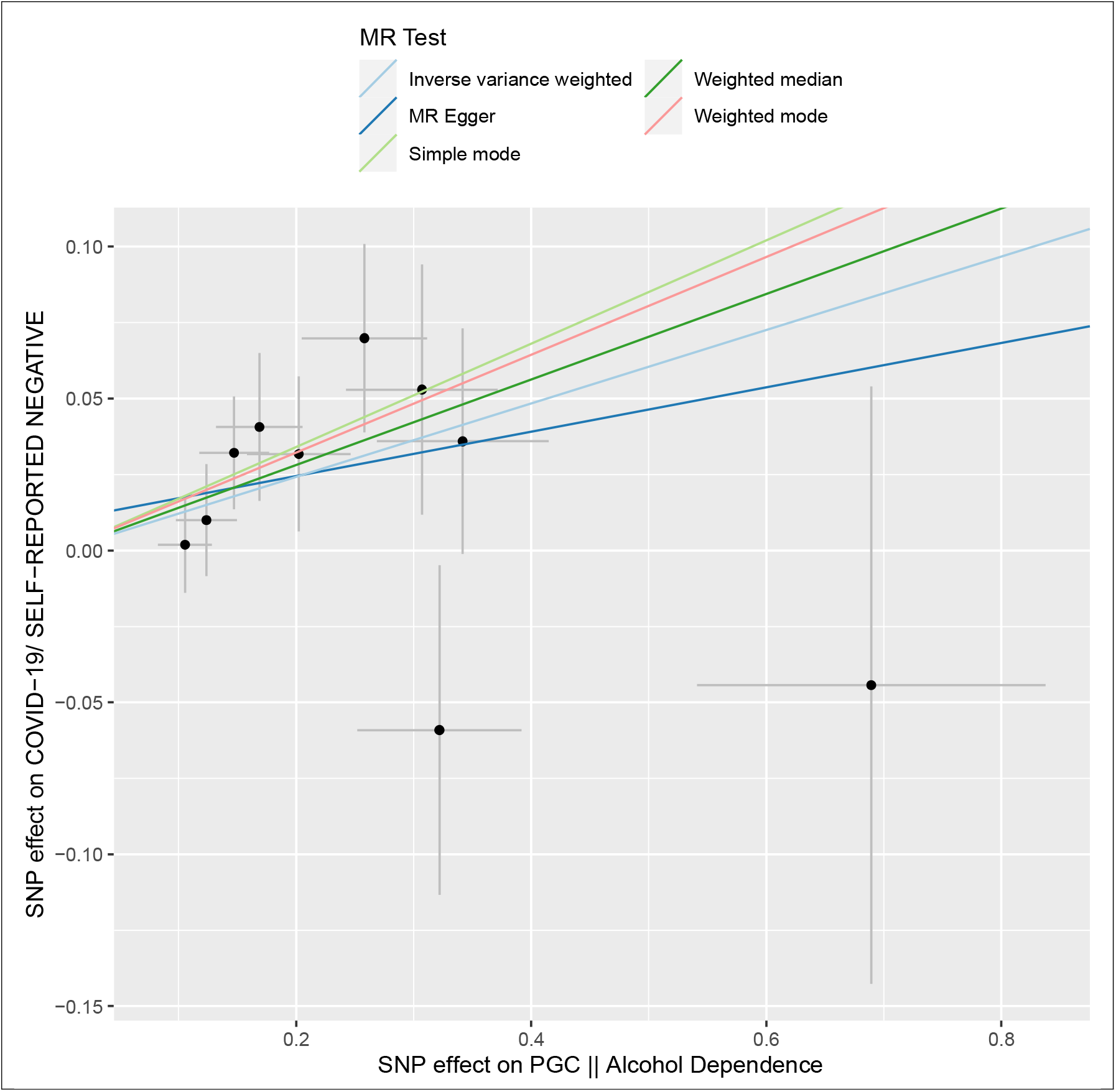
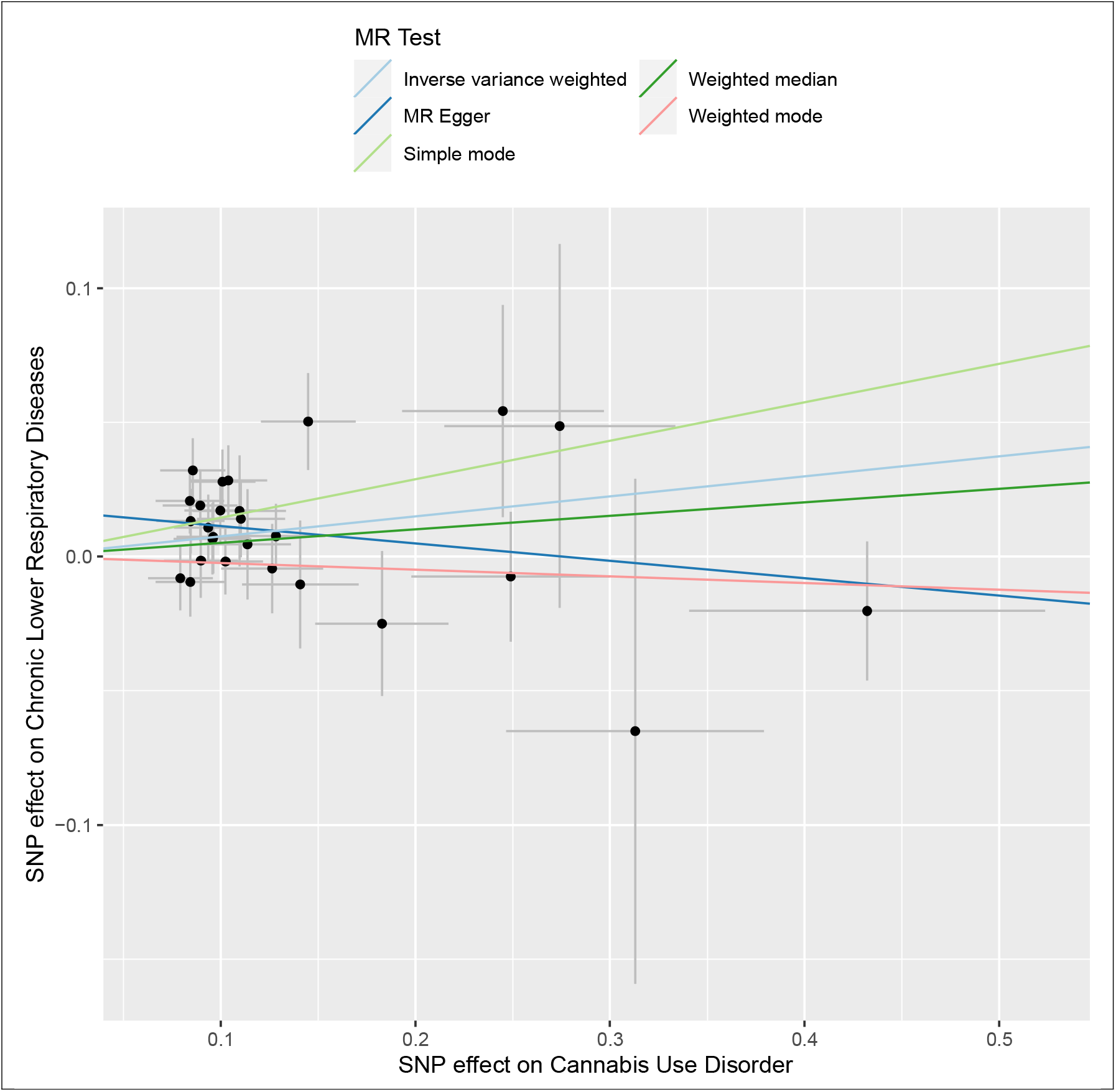
Scatterplot Analysis of Associations of Genetic Liability of Alcohol Dependence on COVID-19 Risk and Cannabis Use Disorder on Chronic Lower Respiratory Disease Risk Scatterplot of single variable Mendelian randomization (SVMR) independent instrument single-nucleotide polymorphism (SNP) exposure effects versus outcome effects from 2 independent samples augmented by the standard error of these effects on the vertical and horizontal sides (for presentation, alleles are coded so that all SNP exposure effects are positive). Solid lines are the regression slopes fitted by the primary inverse variance weighted (IVW) and complementary MR methods: slopes fitted by the IVW MR method were similar in direction and magnitude to slopes fitted by complementary methods. Heterogeneity tests did not indicate residual heterogeneity, and pleiotropy robust methods did not indicate directional bias in these estimates (**Tables 7 and 8 in the <u>Supplement</u>**).

### Tobacco smoking

We included lifetime smoking instruments from the recent GWAS of a lifetime smoking index (which combined smoking initiation, duration, heaviness and cessation), conducted in a sample of 462 690 current, former and never smokers in the UKB (mean sore value 0.359 (S.D. = 0.694); sample, 54 percent female, mean age 56.7 years, 54 percent never smokers, 36 percent former smokers, and 11 percent current smokers).^25^ (An SD increase in lifetime smoking index score would be equivalent to smoking 20 cigarettes per day for 15 years and stopping 17 years previously or 60 cigarettes per day for 13 years and stopping 22 years previously).^25^ For this study, we included all SNPs associated at GWS *P* < 5 × 10^−8^ and clumped at LD *r*^*2*^ = .001 and a distance of 10 000 kb (**Table 2 in the Supplement)**.

**Table 2.** Single Variable MR Results of the Genetic Liability of Alcohol, Cannabis and Lifetime Smoking Exposures on FinnGen Infectious Disease Outcomes

| Outcome | Exposure | N SNPs | Odds Ratio | CI Lower Bound | CI Upper Bound | P-value |
| --- | --- | --- | --- | --- | --- | --- |
| Acute nasopharyngitis (common cold) | Lifetime smoking | 117 | 2.330 | 1.059 | 5.129 | **0.036** |
|  | Cannabis use | 42 | 1.156 | 0.870 | 1.536 | 0.318 |
|  | Cannabis use disorder | 27 | 0.962 | 0.825 | 1.121 | 0.616 |
|  | Drinks per week | 89 | 1.479 | 0.677 | 3.232 | 0.327 |
|  | Alcohol use disorder | 11 | 0.987 | 0.804 | 1.211 | 0.901 |
| Influenza |  |  |  |  |  |  |
|  | Lifetime smoking | 117 | 1.836 | 0.986 | 3.418 | 0.055 |
|  | Cannabis use | 43 | 0.992 | 0.790 | 1.248 | 0.949 |
|  | Cannabis use disorder | 27 | 1.029 | 0.907 | 1.167 | 0.660 |
|  | Drinks per week | 89 | 1.098 | 0.576 | 2.094 | 0.776 |
|  | Alcohol use disorder | 11 | 1.104 | 0.928 | 1.314 | 0.264 |
| Chronic Lower Respiratory Diseases |  |  |  |  |  |  |
|  | Lifetime smoking | 111 | 2.303 | 1.713 | 3.097 | **3.40E-08** |
|  | Cannabis use | 43 | 0.979 | 0.892 | 1.074 | 0.652 |
|  | Cannabis use disorder | 27 | 1.078 | 1.020 | 1.138 | **0.007** |
|  | Drinks per week | 89 | 1.049 | 0.799 | 1.377 | 0.733 |
|  | Alcohol use disorder | 11 | 1.008 | 0.945 | 1.075 | 0.818 |
| Acute Upper Respiratory Infections |  |  |  |  |  |  |
|  | Lifetime smoking | 117 | 1.550 | 1.223 | 1.962 | **2.80E-04** |
|  | Cannabis use | 43 | 1.014 | 0.935 | 1.099 | 0.737 |
|  | Cannabis use disorder | 27 | 1.047 | 0.998 | 1.098 | 0.062 |
|  | Drinks per week | 87 | 1.100 | 0.855 | 1.415 | 0.457 |
|  | Alcohol use disorder | 11 | 0.987 | 0.923 | 1.056 | 0.712 |
| Viral Pneumonia |  |  |  |  |  |  |
|  | Lifetime smoking | 117 | 0.629 | 0.129 | 3.055 | 0.565 |
|  | Cannabis use | 43 | 1.401 | 0.797 | 2.464 | 0.241 |
|  | Cannabis use disorder | 27 | 1.147 | 0.815 | 1.615 | 0.432 |
|  | Drinks per week | 89 | 0.660 | 0.131 | 3.318 | 0.614 |
|  | Alcohol use disorder | 11 | 1.159 | 0.758 | 1.775 | 0.496 |
| Bacterial Pneumoniae |  |  |  |  |  |  |
|  | Lifetime smoking | 117 | 2.083 | 1.352 | 3.208 | **8.70E-04** |
|  | Cannabis use | 43 | 1.083 | 0.933 | 1.256 | 0.296 |
|  | Cannabis use disorder | 27 | 1.092 | 0.998 | 1.194 | 0.056 |
|  | Drinks per week | 89 | 1.023 | 0.659 | 1.587 | 0.921 |
|  | Alcohol use disorder | 11 | 1.034 | 0.904 | 1.182 | 0.628 |
| All Pneumoniae |  |  |  |  |  |  |
|  | Lifetime smoking | 117 | 1.589 | 1.214 | 2.078 | **7.33E-04** |
|  | Cannabis use | 43 | 1.025 | 0.933 | 1.125 | 0.609 |
|  | Cannabis use disorder | 27 | 1.061 | 0.999 | 1.126 | 0.053 |
|  | Drinks per week | 89 | 1.058 | 0.810 | 1.381 | 0.679 |
|  | Alcohol use disorder | 11 | 1.039 | 0.972 | 1.110 | 0.260 |
| Asthma related infections |  |  |  |  |  |  |
|  | Lifetime smoking | 117 | 1.573 | 1.309 | 1.890 | **1.37E-06** |
|  | Cannabis use | 43 | 1.004 | 0.942 | 1.070 | 0.904 |
|  | Cannabis use disorder | 27 | 1.058 | 1.010 | 1.109 | **0.018** |
|  | Drinks per week | 89 | 1.035 | 0.831 | 1.288 | 0.761 |
|  | Alcohol use disorder | 11 | 1.002 | 0.954 | 1.051 | 0.947 |
| Asthma-related acute respiratory infections |  |  |  |  |  |  |
|  | Lifetime smoking | 117 | 1.665 | 1.325 | 2.093 | **1.26E-05** |
|  | Cannabis use | 43 | 0.995 | 0.925 | 1.071 | 0.896 |
|  | Cannabis use disorder | 27 | 1.053 | 1.007 | 1.102 | **0.023** |
|  | Drinks per week | 89 | 1.022 | 0.797 | 1.312 | 0.864 |
|  | Alcohol use disorder | 11 | 1.005 | 0.947 | 1.068 | 0.859 |
| Asthma-related pneumonia or sepsis |  |  |  |  |  |  |
|  | Lifetime smoking | 117 | 1.580 | 1.213 | 2.060 | **7.05E-04** |
|  | Cannabis use | 43 | 1.020 | 0.932 | 1.115 | 0.670 |
|  | Cannabis use disorder | 27 | 1.064 | 1.003 | 1.127 | **0.038** |
|  | Drinks per week | 89 | 1.074 | 0.827 | 1.395 | 0.593 |
|  | Alcohol use disorder | 11 | 1.036 | 0.970 | 1.106 | 0.291 |
| Asthma-related pneumonia |  |  |  |  |  |  |
|  | Lifetime smoking | 117 | 1.662 | 1.276 | 2.165 | **1.67E-04** |
|  | Cannabis use | 43 | 1.025 | 0.936 | 1.123 | 0.596 |
|  | Cannabis use disorder | 27 | 1.070 | 1.007 | 1.137 | **0.028** |
|  | Drinks per week | 89 | 1.076 | 0.818 | 1.414 | 0.601 |
|  | Alcohol use disorder | 11 | 1.033 | 0.966 | 1.104 | 0.339 |
| Asthma/COPD (Kela code 203) | Lifetime smoking | 112 | 2.069 | 1.471 | 2.909 | **2.90E-05** |
|  | Cannabis use | 42 | 0.966 | 0.845 | 1.105 | 0.613 |
|  | Cannabis use disorder | 27 | 1.048 | 0.985 | 1.114 | 0.136 |
|  | Drinks per week | 89 | 1.059 | 0.775 | 1.447 | 0.720 |
|  | Alcohol use disorder | 11 | 1.014 | 0.935 | 1.099 | 0.745 |

Abbreviations: MR, mendelian randomization; GWAS, genome wide association study; N SNPs, number of single-nucleotide polymorphism (genetic instruments); OR, odds ratio; CI, confidence interval. Boldface indicates conventional statistical significance ( $P < 0.05$ ).
a Results from two sample SVMR inverse-variance weighted MR analysis; outliers identified by MR PRESSO tool were removed; estimated associations reported as odds ratios with 95% confidence intervals.
b Genetic instruments selected from 5 GWASs, selection threshold $P < 5 \times 10^{-8}$ or $P < 5 \times 10^{-6}$ , clumped at linkage disequilibrium (LD) $r^2 = .001$ (10 000 kilobase pair window); N SNPs differs across outcomes depending on number of genetic instruments found in outcome GWASs.

### Cannabis use

We included two cannabis-related instrument sets: cannabis use and CUD. Summary statistics for lifetime cannabis use (a yes/no variable of whether participants reported using cannabis during their lifetime) were obtained from the PGC meta-analysis GWAS of 3 cohorts (International Cannabis Consortium (35 297 respondents, 55.5 percent female, ages 16-87, mean 35.7 years; 42.8 percent had used cannabis); UKB (126 785 respondents, 56.3 percent female, aged 39-72, mean age 55.0 years, 22.3 percent had used cannabis), and 23andMe (22 683 respondents, 55.3 percent female, ages 18-94, mean 54.0 years; 43.2 percent had used cannabis)).^26^ CUD instruments were obtained from a recent PGC meta-analysis of 3 cohorts of predominantly European ancestry (PGC, Lundbeck Foundation Initiative for Integrative Psychiatric Research (iPSYCH), and deCODE, excluding related individuals from PGC family-based cohorts; demographics not available), including 14 808 cases of cannabis abuse or dependence defined as meeting DSM-IIIR, DSM-IV, DSM-5, or ICD10 codes (depending on study cohort) criteria; the 358 534 controls were defined as anyone not meeting the criteria.^27^ To ensure independence, we included all SNPs associated with cannabis use at GWS *P* < 5 × 10^−8^ and all SNPs association with CUD at *P* < 5 × 10^−6^ (due to absence of GWS SNPs^28-30^), clumped, respectively, at LD *r*^*2*^ = .001 and a distance of 10 000 kb (**Table 3 in the Supplement)**.

### Alcohol consumption

We included three instrument sets related to alcohol use: drinks per week,^31^ frequency of binge drinking (consuming six or more units of alcohol per occasion),^32^ and AUD. Drinks per week instruments were obtained from the GSCAN GWAS meta-analysis of 29 cohorts (941 280 individuals; demographics not available) of predominantly white European ancestry.^31^ Given the varied cohort methods used to measure alcohol consumption (binned, normalized, etc.), the data was log transformed: thus, the effect estimate is measured in log transformed drinks per week.^31^ To ensure independence, we included all SNPs associated at conventional genome-wide significance (GWS) (*P* < 5 × 10^−8^) and clumped at linkage disequilibrium (LD) *r*^*2*^ = .001 and a distance of 10 000 kilobase (kb) (**Table 4 in the Supplement)**.

For the instrument set assessing frequency of consuming six or more drinks (binge drinking), we used Alcohol Use Disorder Inventory Test (AUDIT) question 3 (“How often do you have six or more drinks on one occasion?”) derived from the Neale Lab GWAS results from 143 658 UKB participants of European ancestry.^32^ For the AUD instrument set, we used the Psychiatric Genomics Consortium (PGC) GWAS meta-analysis of 28 cohorts (51.6 percent female, 8 485 cases, 20 657 controls) of predominantly European ancestry.^33^ AUD was diagnosed by either clinician rating or semi-structured interview using DSM-IV criteria including the presence of at least three of seven alcohol-related symptoms (withdrawal, drinking larger amounts/drinking for longer time; tolerance; desire or attempts to cut down drinking; giving up important activities to drink; time related to drinking; or continued alcohol consumption despite psychological and/or physical problems^34^). To ensure independence, we included all SNPs associated at *P* < 5 × 10^−6^ (due to absence of GWS SNPs^28-30^) and clumped at LD *r*^*2*^ = .001 and a distance of 10 000 kb (**Table 2 in the Supplement)**.

For the multivariable MR (MVMR) analyses, we concatenated independent instrument sets for alcohol use, cannabis use and lifetime smoking, and AUD, CUD, and lifetime smoking (clumping the resulting two MV instrument sets to exclude intercorrelated SNPs with pairwise LD *r*^*2*^ > .001), giving us 141 and 126 MV instruments, respectively. *F* statistics for the unconditional instruments were strong (> 10, **Tables 3-5 in the Supplement**). We were unable to calculate conditional *F* statistics to assess the strength of the multivariable instrument sets: SVMR statistical methods recently extended to two sample MVMR are appropriate only for non-overlapping exposure summary level data sources; when overlapping, the requisite pairwise covariances between SNP associations are determinable only using individual level data.^35^

### COVID-19 outcomes

We used summary GWAS statistics from the COVID-19 Host Genetics Initiative (COVID-19 hg) meta-analysis Round 4 release data (20 October 2020) (https://www.covid19hg.org/results)^36^ for 9 COVID-19 phenotypes in predominantly European ancestry cohorts (COVID-19 HG coding; N cases; N controls; demographics not available): very severe respiratory confirmed COVID-19 versus population (A2_ALL; 4 933; 623 902); very hospitalized COVID-19 versus not hospitalized COVID-19 (B1_ALL; 2 430; 8 478); hospitalized COVID-19 versus population (B2_ALL; 7 885; 961 804); very severe hospitalized COVID-19 versus population European cohorts only (B2_ALL_EUR; 6 406; 902 088); COVID-19 versus lab/self-reported negative COVID-19 test (C1; 11 085; 116 794); COVID-19 versus population (C2_ALL; 17 965; 1 370 547); COVID-19 versus population European ancestry only (C2_ALL_EUR; 14 134; 1 284 876); and predicted COVID-19 from self-reported symptoms versus predicted/self-reported negative (D1_ALL, 3 204; 35 728) (**Table 1 in the Supplement**).

### Other respiratory infection and disease outcomes

We used data from FinnGen project Datafreeze 3 for additional respiratory-related outcomes (**Table 1 in the Supplement**)^37^. Detailed documentation is provided on the FinnGen study website (https://finngen.gitbook.io/documentation/). FinnGen is a public-private partnership incorporating genetic data for disease endpoints from Finnish biobanks and Finnish health registry EHRs.^37^ FinnGen Datafreeze 3 samples included only European ancestry participants; samples sizes ranged from 116 335 (acute nasopharyngitis) to 135 638 (acute URIs, asthma-related infections, asthma-related pneumonia or sepsis, bronchitis, chronic LRIs, and influenza and pneumonia) (**Figure 1; Table 1 in the Supplement**).

### Sample independence

Participant overlap in samples used to estimate genetic associations between exposures and outcomes can increase weak instrument bias (WIB) in MR analyses,^38,39^ but to a lesser extent with large biobank samples (including UKB and deCODE).^40^ Given the large size of the overlapping UKB and deCODE cohorts (**Table 1 in the Supplement**) and the strength of the instruments in both directions (*F* statistics > 10; **Tables 2-4 in the Supplement**), considerable WIB would not be expected.^39,40^

### Statistical and sensitivity analyses

For SVMR analyses, we used inverse-variance weighted MR (MR IVW) along with MR-Egger, weighted median, and weighted mode methods, to assess evidence of causal effects of each of alcohol, cannabis and tobacco use and dependence on infectious disease outcomes; also, to detect the sensitivity of the results to different patterns of violations of IV assumptions,^41^ as consistency of results across methods strengthens an inference of causality.^41^ For MVMR analyses, we used an extension of the IVW MR method, performing multivariable weighted linear regression (variants uncorrelated, random-effects model) with the intercept term set to zero.^38,42^ We used an extension of the MR-Egger method to correct for both measured and unmeasured pleiotropy ^43^. To evaluate heterogeneity in instrument effects, which may indicate potential violations of the IV assumptions underlying two-sample MR.^44^ we used the MR Egger intercept test,^44^ and the Cochran heterogeneity test,^45^ and multivariable extensions thereof.^42,43^ The MR <u>p</u>leiotropy <u>res</u>idual <u>s</u>um and <u>o</u>utlier (MR-PRESSO) global test, and multivariable extension thereof ^46^, were used to facilitate identification and removal of outlier instruments to correct potential directional horizontal pleiotropy and resolve detected heterogeneity. For the SVMR, we used the Steiger directionality test tested the causal direction between the hypothesized exposure and outcomes.^47^ Analyses were carried out using TwoSampleMR, version 0.5.5,^41^ MendelianRandomization, version 0.5.0, and MR PRESSO, version 1.0,^46^ in the R environment, version 4.0.2.

### Reported results and interpretation of findings

SVMR and MVMR results with test statistics both before and after outlier correction are presented in **Tables 7-11 in the Supplement**. MR IVW (MR PRESSO outlier corrected) odds ratios (OR) with 95% CI, per unit increase in the exposures (*e*.*g*. per unit increase of log-transformed alcoholic drinks per week or lifetime smoking index), with *P*-value, are presented in **Tables 1-2**.

While we caution against interpreting the results based solely upon a *P*-value threshold,^48^ we generally use a two-sided α of .007 based on comparing 7 COVID-19 outcomes and .004 based on comparing14 FinnGen infectious respiratory diseases as a heuristic that allows for follow-up analyses for a plausible number of findings. In assessing consistency and robustness, we looked for estimates substantially agreeing in direction and magnitude (overlapping confidence intervals) across then four complementary MR methods used. We underscore evidence strength based upon the effect magnitude and direction, the 95% confidence interval of that effect, and the *P-*value.

## RESULTS

### COVID-19 disease outcomes

In SVMR, genetically predicted lifetime smoking was found to be associated with increased risk for “hospitalized COVID-19 versus population” (OR 4.039, 95% CI 2.335-6.985, *P*-value = 5.93×10^−7^) and “very severe hospitalized COVID-19 versus population (predominantly European)” (OR 3.091, 95% CI 1.883-5.092, *P*-value = 8.40×10^−6^), with consistent magnitude and direction across all four SVMR methods (**Table 1; Table 8 in the Supplement**). Controlling for AUD and CUD (**Table 1; Table 10 in the Supplement**), and alternatively for alcohol and cannabis use (**Table 1; Table 11 in the Supplement**), in MVMR, genetically predicted lifetime smoking was still found to be associated with increased risk for both COVID-19 outcomes, with increased point estimate magnitudes compared to SVMR (“hospitalized COVID-19 versus population” OR 4.457, 95% CI 2.287-8.687, *P*-value = 1.14×10^−5^ (controlling for other substance dependence/use disorder), and OR 4.716, 95% CI 2.580-8.621, *P*-value = 4.66×10^−7^ (controlling for other substance use); and “very severe hospitalized COVID-19 versus population (predominantly European)”, OR 3.152, 95% CI 1.726-5.757, *P*-value = 1.86×10^−4^ (controlling for other substance dependence/use disorder), and OR 3.630, 95% CI 2.126-6.199, *P*-value = 2.33×10^−6^ (controlling for other substance use)), with consistent magnitude and direction across the two MVMR methods.

Genetically predicted AUD but not alcohol consumption (drinks per week) was found to be associated with increased risk for “COVID-19 versus self-reported negative” in SVMR (OR 1.128, 95% CI 1.039-1.226, *P*-value = .004); in MVMR, controlling for cannabis use disorder and lifetime smoking, that association remained although with a lesser *P*-value (OR 1.145, 95% CI 1.033-1.269, *P*-value = .010) (**Tables 8 and 10 in the Supplement**). In contrast, genetically predicted cannabis use and CUD were not found to be associated with risk of any COVID-19 disease outcome (**Tables 7 and 9-11 in the Supplement**).

### FinnGen infectious disease outcomes

In SVMR, genetically predicted lifetime smoking was found to be associated with increased risk for: Bronchitis (OR 1.558, 95% CI 1.196-2.030, *P*-value = .001), all pneumoniae (OR 1.589, 95% CI 1.214-2.078, *P*-value = 7.33×10^−4^), bacterial pneumonia (OR 2.083, 95% CI 1.352-3.208, *P*-value = 8.70×10^−4^), acute upper respiratory infections (OR 1.550, 95% CI 1.223-1.962, *P*-value = 2.80×10^−4^, chronic lower respiratory diseases (OR 2.303, 95% CI 1.713-3.097, *P*-value = 3.40×10^−8^), asthma related infections (OR 1.573, 95% CI 1.309-1.890, *P*-value = 1.37×10^−6^), asthma related acute respiratory infections (OR 1.665, 95% CI 1.325-2.093, *P*-value = 1.26×10^−5^), asthma-related pneumonia (OR 1.589, 95% CI 1.214-2.078, *P*-value = 1.67×10^−4^), asthma/ COPD (OR 2.069, 95% CI 1.471-2.909, *P*-value = 2.90×10^−5^), and asthma-related pneumonia or sepsis (OR 1.580, 95% CI 1.213-2.060, *P*-value = 7.05×10^−4^). In MVMR, controlling for AUD and CUD, and, alternatively, for alcohol and cannabis use, lifetime smoking was still found to be associated with increased risk for these outcomes, with consistent magnitude and direction across SVMR and MVMR outcomes but with *P*-values attenuated towards the null ((**Tables 10-11 in the Supplement**)).

Genetically predicted CUD, but not cannabis use, was found associated with increased risk for chronic lower respiratory diseases (including bronchitis and asthma COPD) (OR 1.078, 95% CI 1.020-1.128, *P*-value = .007); bronchitis (OR 1.061, 95% CI 1.003-1.124, *P*-value = .040); asthma related acute respiratory infections (OR 1.053, 95% CI 1.007-1.102, *P*-value = .023); asthma related pneumonia (OR 1.070, 95% CI 1.007-1.137, *P*-value = .028); and asthma related sepsis or pneumonia (OR 1.064, 95% CI 1.003-1.127, *P*-value = .038) (**Table 9 in the Supplement**). Controlling for AUD and lifetime smoking in MVMR, these associations attenuated towards the null (**Table 10 in the Supplement**). In contrast, genetically predicted alcohol use and AUD were not found to be associated with risk of any FinnGen infectious disease outcome (**Tables 7 and 9-11 in the Supplement**).

Evidence of heterogeneity but not directional pleiotropy was found only for lifetime smoking on chronic lower respiratory diseases across SVMR and MVMR analyses; and also for lifetime smoking on asthma/COPD in the MVMR analyses controlling for AUD and CUD. Steiger directionality analyses suggests correct causal direction for all analyses (**Tables 7-11 in the Supplement**).

## DISCUSSION

Using large summary-level GWAS data and complementary two-sample MR methods, we show that the genetic liability for tobacco smoking has potential causal relationships with several respiratory infection and disease outcomes, including COVID-19. These tobacco smoking-respiratory findings were supported by multivariable MR analyses accounting for alcohol and cannabis use and abuse, which in addition the broadly consistent IVW results (within the IVW MR 95% confidence interval but typically less precise) with estimates from the weighted median, weighted mode, and MR Egger sensitivity analyses strengthens causal inference. Further, in single variable MR, we identify potential adverse impact CUD on lower respiratory infection, the common cold, and several asthma-related infections, suggesting evidence for a dose-dependent impact of cannabis use where heavy cannabis use may be harmful to the respiratory system. In parallel, we find little evidence for an alcohol-respiratory infection relationship suggesting that previous observational data may be due to confounding.

Our COVID-19 results extend recent MR studies showing adverse effects of smoking on COVID-19 risk^49^ by accounting for highly comorbid alcohol consumption, cannabis use, and SUDs, which when combined with reports suggesting smoking intensifies the severity of COVID-19 symptoms,^50,51^ the risk for being admitted to an intensive care unit or requiring ventilation,^51^ and recent transcriptomics-based work showing that smoking may increase the expression of angiotensin converting enzyme 2 (ACE2), the putative receptor for severe acute respiratory syndrome coronavirus 2 (SARS-CoV-2) (the virus that causes COVID-19),^52^ suggests smoking may be an important modifiable risk factor for COVID-19 risk.

Our genetics-based findings support and extend the observational literature identifying tobacco smoking as a risk factor for respiratory infection and diseases,^9,53,54^ and add to the recent MR literature identifying potential causal links of smoking with reduced lung function^55^, lung cancer,^56^ and increased mortality due to respiratory disease.^57^ Potential mechanisms by which smoking increases respiratory infection risk include structural changes to the respiratory tract and a dysregulated cellular and humoral immune response, including peribronchiolar inflammation, decreased levels of circulating immunoglobulins, and changes to pathogen adherence.^53^ For example, smoking has been shown to stimulate the release of catecholamine and corticosteroids, which may, in turn, increase circulating CD8^+^ lymphocytes and suppress the host defense against infections.^53^ Notably, many immunological effects related to smoking may resolve within six weeks of smoking cessation,^53^ which suggests that smoking cessation programs may have an important impact on reducing respiratory infections.

Regarding cannabis use, to our knowledge this is the first MR study to investigate the role of cannabis use in respiratory infections, and while we failed to find evidence of any relationships, smoking cannabis, like tobacco smoking, may prompt the onset of coughing, which could consequently increase viral transmission, or may exacerbate possibly respiratory symptoms. As cannabis is the most used drug worldwide –an estimated 188 million recreational users worldwide^58^ – this aspect of cannabis use may have important implications for the spread of COVID-19. In contrast, the single-variable MR CUD results demonstrated adverse effects on several respiratory outcomes, but not COPD, which supports the existing literature;^59-61^ however, accounting for lifetime tobacco smoking attenuated highlighting the complex nature of these relationships. Further, habitual cannabis smoking may have several effects on respiratory and immune systems that may impact respiratory infection susceptibility: For example, structural abnormalities in alveolar macrophages and coincident dysregulated cytokine production and antimicrobial activity have been reported.^61^ While our study provides preliminary genetic evidence suggesting potential causal relationships between heavy cannabis use and respiratory infection, additional triangulating lines of evidence (i.e., immune monitoring studies), are required to further elucidate the CUD-respiratory infection relationship. However, the toxin and irritant profiles of cannabis and tobacco smoke are similar,^11^ which suggests the direct route of administration via inhalation for these substances and may result in dysregulated pulmonary physiology which may, in turn, increase infection risk.

In contrast to our tobacco smoking findings, we failed to find genetic evidence of respiratory implications due to alcohol consumption not meeting threshold of AUD, or binge drinking, suggesting that previous observational literature may be due to confounding from other comorbid behaviors – such as smoking – that may be the true causal risk factors for respiratory infections. For example, observational and genetic evidence have shown a strong association between alcohol consumption and smoking: It has been estimated that 85% of smokers consume alcohol^62-64^ and alcohol drinkers are 75% more likely than abstainers to smoke^65^. Therefore, it is possible that the observational study-based alcohol-respiratory infection links may be due, instead, to tobacco smoking; however, future work will be needed to confirm this hypothesis. In addition, it is important to note that our results should not be interpreted as suggesting that alcohol does not impact overall lung health and structure, which has been previously reported.^7^ Further, while we failed to find evidence that weekly alcohol consumption impacted COVID-19 risk, the Centers for Disease Control recently showed that dining at on-site locations, such as restaurants and bars, is associated with increased COVID-19 risk,^66^ and given that alcohol consumption may lower inhibition and increase impulsivity,^67^ individuals consuming alcohol may take social distancing less seriously, and thereby unintentionally spread the SARS-CoV-2 virus.

This study has several strengths including the use of multiple alcohol consumption and cannabis use variables, which enabled us to evaluate various dimensions of substance use and abuse and identify possible causal relationships of substance use disorders and respiratory outcomes. In addition, our main single variable analyses included multiple MR methods, each relying on orthogonal assumptions, provides confidence in robustness of the results and strengthens causal inference.^68^ Further, our multivariable two-sample MR design, the most appropriate design given the strong correlation between tobacco smoking, alcohol consumption and cannabis use, yielding estimates that account for these correlated behaviors for each exposure on COVID-19 risk and other respiratory outcomes.^69^

This study also has limitations: For example, like existing self-reported substance use literature, these exposures may be either under- or over-reported.^70^ Because many of the datasets included UK Biobank participants, who are more educated, with healthier lifestyles, and fewer health problems than the UK population,^71^ which may limit the applicability of our findings to other populations. Regarding our mainly null alcohol-respiratory infection results, it is possible that alcohol may have indirect impact on infection risk through a modified immune response,^72^ or other system dysregulation, that may modulate infection risk that we were not able to directly assess. Further, while we found some evidence that AUD may increase the risk for COVID-19; the largely null other current AUD findings does not support a broader AUD-respiratory disease relationship. However, like other recent psychiatric MR studies where the exposure instruments included a relaxed statistical threshold, our binge drinking and AUD instruments were comprised of independent SNPs associated with the respective drinking behavior (i.e., P-value < 5×10^−6^) for SNP inclusion due to the lack of conventionally GWS SNPs (P-value < 5×10^−8^),^29,30^ which may impact the results. Because heavy alcohol consumption and AUD have been previously linked with acute respiratory distress syndrome^10^ – one of the most severe complications of COVID-19,^73^ future studies re-evaluating the links between heavy alcohol consumption and AUD when better powered GWAS data becomes available.

In addition, the included samples were comprised of primarily white individuals of European ancestry, and research has shown strong racial, ethnic, and socioeconomic disparities in COVID-19 risk, and severity.^74-76^ Therefore, we caution the generalization of these findings and urge future work to investigate these relationships using a genetics-based approach in other populations when the data becomes available. Another limitation is the overlap of the UKB participants between the alcohol consumption, lifetime smoking, and COVID-19 outcomes, which may bias resulting estimates;^39^ however, any bias would likely be minimal.^39^ It has been also shown that two-sample MR may be safely used in single samples provided the data is derived from large biobanks (i.e., the UKB, FinnGen, etc.,).^77^

In conclusion, our data provide genetic evidence of adverse relationships between smoking and many respiratory-related disease outcomes ranging from the common cold to severe COVID-19, which suggests prevention programs aimed at smoking cessation and prevention may have public health and clinical benefits. We also observed a potential dose-dependent relationship where heavy substance use – as indicated by CUD – but not general cannabis use, was linked with several respiratory infections, suggesting heavy cannabis use may have a harmful impact on respiratory infections, which may have important respiratory-related consequences given the increasingly permissive cannabis-use laws.

## Data Availability

All analyses were based upon publicly available data. COVID-19 GWAS summary-level data is available at https://www.covid19hg.org/results/. FinnGen data are available at https://www.finngen.fi/en; lifetime smoking at https://data.bris.ac.uk/data/dataset/10i96zb8gm0j81yz0q6ztei23d; alcohol drinks per week data at: https://genome.psych.umn.edu/index.php/GSCAN; frequency of consuming >6 drinks per occasion data are available at: http://www.nealelab.is/uk-biobank; cannabis use disorder and alcohol dependence data are available through the Psychiatric Genomics Consortium data portal: https://www.med.unc.edu/pgc/download-results/; and the cannabis use data are available through the International Cannabis Consortium at: https://www.ru.nl/bsi/research/group-pages/substance-use-addiction-food-saf/vm-saf/genetics/international-cannabis-consortium-icc/.

## ACKNOWLEGEMENTS

We want to acknowledge the participants and investigators of the many studies used in this research without whom this effort would not be possible: the COVID-19 Host Genetics Initiative and the contributors thereto specified at http://www.covid19hg.org/acknowledgments.html; FinnGen study; and the UK Biobank. We also want to acknowledge the Medical Research Council Integrative Epidemiology Unit (MRC-IEU, University of Bristol, UK), especially the developers of the MRC-IEU UK Biobank GWAS Pipeline. This work was supported by the National Institutes of Health (NIH) intramural funding [ZIA-AA000242 to F.W.L]; Division of Intramural Clinical and Biological Research of the National Institute on Alcohol Abuse and Alcoholism (NIAAA).

